# Elucidating Delays in Illness Recognition, Healthcare Seeking, and Healthcare Provision for Stillbirths and Neonatal Deaths in Seven Low- and Middle-Income Countries

**DOI:** 10.1101/2025.10.07.25337535

**Authors:** Kitiezo Aggrey Igunza, Namrata Ramakrishna, Zachary J. Madewell, Victor Akelo, Hellen Muttai, Christopher Mugah, Paul Mitei, Ikechukwu Ogbuanu, Ima-Abasi Bassey, Erick Kaluma, Oluseyi Balogun, Ronita Luke, Shams El Arifeen, Emily S. Gurley, Mohammad Zahid Hossain, Rajib Biswas, Afsana Afrin, Nega Assefa, J. Anthony G. Scott, Haleluya Leulseged, Meron Kebede, Konjit Eshetu, Karen L. Kotloff, Samba O. Sow, Rosauro Varo, Inácio Mandomando, Marcelino Garrine, Milton Kincardett, Quique Bassat, Shabir A. Madhi, Ziyaad Dangor, Megan Dempster, Marguerite Hall, Sithembiso Velaphi, John Blevins, Maria Maixenchs, Yuting Guo, Abeed Sarker, Portia C. Mutevedzi, Cynthia G. Whitney, Chris A. Rees, Child Health and Mortality Prevention Surveillance (CHAMPS) Network

## Abstract

An estimated 1.9 million stillbirths and 2.3 million neonatal deaths occurred globally in 2023, disproportionately clustered in sub-Saharan Africa and South Asia. We aimed to determine the frequency of delays in illness recognition, healthcare seeking, and healthcare receipt among cases of stillbirth neonatal death, to characterize the most common delays, and to describe the frequency of antenatal care visits preceding stillbirths and neonatal deaths. We conducted a descriptive analysis of deaths enrolled during 2016-2023 in the Child Health and Mortality Prevention Surveillance (CHAMPS) network, which prospectively studies stillbirths and child deaths in seven low- and middle-income countries. Delays in 1) illness recognition in the home/decision to seek care, 2) transportation to healthcare facilities, and 3) the provision of clinical care (e.g., timely administration of medications, etc.) in healthcare facilities were categorized according to the “Three Delays-in-Healthcare” framework. We included 7,079 deaths (3,424 stillbirths and 3,655 neonatal deaths). Among included stillbirths, 53.6% were male and the median maternal gestational age was 36 weeks (interquartile range [IQR] 32, 38 weeks). Among included neonatal deaths, 57.3% were male and the median maternal gestational age at delivery was 34 weeks (IQR 30, 38 weeks). Overall, 96.8% (n=3,313) of stillbirths and 97.7% (n=3,571) of neonatal deaths had ≥1 delay identified, the most common being those related to the delivery of high-quality clinical care in health facilities, despite 84.9% of stillbirths and 82.9% of neonatal deaths occurring in healthcare facilities. Nearly half of mothers of stillbirths (47%) and 41% of mothers of neonates who died had ≤3 antenatal care visits recorded. Despite high facility delivery rates, healthcare provision delays were the most predominant, suggesting that strengthening facility-based clinical care quality, rather than improving facility access, may be the most impactful intervention for reducing perinatal mortality in these settings.

## Introduction

Despite massive reductions in mortality among young children aged <5 years from the 1990s to the 2020s, declines in stillbirth and neonatal mortality rates have been more modest.^1^ Worldwide, there were an estimated 1.9 million stillbirths and approximately 2.3 million neonatal deaths in 2023.^2,3^ Sub-Saharan Africa and South Asia, where 56% of all live births occur, account for >70% of global stillbirths and >80% of global neonatal deaths.^4,5^

Results from prior studies suggest that delayed decisions to seek healthcare by caregivers, difficulty accessing healthcare facilities, and suboptimal clinical care in healthcare facilities may contribute to high rates of childhood mortality in sub-Saharan Africa and South Asia.^6–9^ Many stillbirths and neonatal deaths are thought to be preventable when mothers access skilled healthcare personnel in a timely fashion.^2,3^ We have previously applied the “Three Delays in Healthcare” framework to better understand social (i.e., identification of illness in the home and transportation to healthcare facilities) and health system (i.e., adequacy and quality of clinical care provided) issues contributing to mortality among infants and children aged 1 to 59 months in Bangladesh and six sub-Saharan African countries.^10^ Stillbirths and neonatal deaths were not included in those analyses because of anticipated differences in healthcare seeking for these populations, the interconnectedness of pregnancy-related illnesses with stillbirths and neonatal outcomes, and differences in health services delivery for neonates compared to young children.^11^ Here, we provide insights to better understand healthcare seeking and provision factors that may contribute to persistently high rates of stillbirths and neonatal deaths in sub-Saharan Africa and South Asia.^12,13^

A detailed understanding of the most common delays encountered by caregivers of stillbirths and neonates who died has the potential to inform both social and clinical interventions tailored to prevent future stillbirths and neonatal deaths. Our objectives were 1) to determine the frequency of delays among stillbirths and neonatal deaths in the Child Health and Mortality Prevention Surveillance (CHAMPS) network, 2) to characterize the most common delays within the “Three Delays in Healthcare” framework, and, 3) because clinical care that may impact perinatal survival begins during pregnancy, to describe the frequency and timing of antenatal care visits among mothers of stillbirths and neonatal deaths.

## Materials and Methods

### Study Design

We conducted a descriptive analysis using data collected in CHAMPS, a prospective stillbirth and child mortality surveillance network.^14,15^ CHAMPS tracks stillbirths and deaths among children aged <5 years and performs postmortem testing to determine underlying, immediate, and co-morbid causes of death for all enrolled cases. Additionally, CHAMPS determines whether the stillbirth or neonatal death could have been prevented and provides insights into possible interventions that could have prevented the deaths. Ethical clearance for CHAMPS was granted by ethical review boards from each participating site and by Emory University Rollins School of Public Health.

### Study Setting

CHAMPS operates in regions with disproportionately high rates of stillbirths and childhood deaths at sites in seven countries in South Asia and sub-Saharan Africa (i.e., Baliakandi and Faridpur, Bangladesh; Kersa, Haramaya, and Harar, Ethiopia; Kisumu and Siaya, Kenya; Bamako, Mali; Manhiça and Quelimane, Mozambique; Makeni and Bo, Sierra Leone; and Soweto, South Africa).^16^ At each site, CHAMPS staff conduct mortality surveillance in both the community (i.e., at the level of the household) and within healthcare facilities (i.e., outpatient clinics, community hospitals, and referral hospitals).

### Population Inclusion and Exclusion Criteria

We included stillbirths (weighing ≥1,000 grams or born at ≥28 weeks gestation) and live-born babies aged 0 to 27 days at the time of death (neonatal deaths) that enrolled in CHAMPS from June 15, 2016 to December 31, 2023. For this analysis, we excluded deaths among infants and young children aged 1-59 months enrolled in CHAMPS because delays in healthcare seeking and provision among these populations in CHAMPS have been described previously.^10^ CHAMPS enrolls stillbirths and neonatal deaths in two ways. First, cases in which the family of the deceased provides written informed consent for the deceased to undergo full enrollment that includes extensive extraction of clinical data, verbal autopsy, postmortem testing, and determination of cause of death by an expert panel.^17,18^ For cases in which families do not consent to postmortem testing, data from clinical records and verbal autopsies are still collected, but no complete cause of death determination is conducted. As our aim was to understand delayed illness recognition by caregivers, transport to healthcare facilities, and delays in healthcare facilities, we included both types of CHAMPS-enrolled cases because requisite data for these analyses were available for both types of cases. However, our analyses regarding delays stratified by causes of death were limited to those cases that underwent complete postmortem testing.

### Data Sources

For each enrolled stillbirth or neonatal death, CHAMPS staff conducted verbal autopsies in accordance with the World Health Organization (WHO) Verbal Autopsy form.^19^ This form contains both discrete and open-ended questions about signs and symptoms for the deceased before death as well as questions about antemortem healthcare seeking behaviors. CHAMPS staff also inquire about antenatal care and maternal illnesses for mothers of stillbirths and neonatal deaths. Additionally, for each enrolled case in CHAMPS, staff conduct extensive review of all available antemortem clinical records (i.e., clinic notes, hospital admission forms, progress notes, laboratory and radiology results, etc.) for the mother, stillbirth (if records are available), and the deceased neonate. Data from clinical charts are recorded in CHAMPS in both discrete and narrative summary form, making note of both the clinical care that was provided as well as any deficits in clinical care provided. Narrative and discrete variables collected in CHAMPS were used to identify delays that occurred before death according to the “Three Delays in Healthcare” framework.

### Variables

Prior to any data extraction, we developed a list of variables that represented each delay in the “Three Delays-in-Healthcare” framework after reviewing variables identified in prior studies on illness recognition in the home/decision to seek care (Delay 1), delays in identifying and transportation to a healthcare facility (Delay 2), and delays in the provision of quality clinical care (Delay 3).^6–9^ We then piloted the use of our list of potential delays through reading narrative fields and discrete variables of 100 randomly-selected cases across the included sites. During that phase, we iteratively added additional delays that were not part of the initial list of potential delays related to antenatal care, maternal conditions, illness recognition in the home, reaching a healthcare facility, and the provision of clinical care. Facility deaths are defined in CHAMPS as those that occur in a healthcare facility. Community deaths are considered those that occur outside formal healthcare settings.

### Data Extraction

Two reviewers independently read through narrative data fields from verbal autopsies and clinical summaries for ∼20% (n=864) of all cases to extract data on delays identified in text fields. We then experimented with two state-of-the-art open-source large language models (LLAMA-3-70B and Mixtral 8×7B)^20,21^ to assess their performance compared to the results of our manual review as a reference standard. We selected the large language model that demonstrated an initial precision of >0.95 and an F_1_ score >0.95 (i.e., LLAMA-3-70B). In the training phase, we ran the model using the 20% of cases that were reviewed by two reviewers and reviewed any discrepancies between the model’s output and manual review. We fine-tuned the model prompt based on patterns observed in discrepancies caused by model errors. The optimized LLAMA-3-70B model was then used to extract data on identified delays among the remaining 80% of cases. A subset of the remaining 80% of cases that were not reviewed by the two reviewers initially was then reviewed to identify any additional discrepancies between the model’s output and manual review. This review again resulted in a precision of >0.95 and an F_1_ score >0.95, so data generated from the LLAMA-3-70B model were used for our analyses.

### Analyses

We used descriptive statistics for demographics of enrolled cases and to determine the frequency of each delay type with the “Three Delays in Healthcare” framework. Based on prior evidence that suggests delays in healthcare seeking may vary by the sex of a child and by site, we compared the frequency of delays by these factors.^10,22–24^ We also plotted the frequency of each class of delay by year to assess trends over time. Furthermore, we assessed whether delays within the “Three Delays in Healthcare” framework were more common among mothers with certain maternal conditions (e.g., hypertension, etc.). We used chi-square tests to determine whether delays varied by cases enrolled at different sites within the CHAMPS network, by sex, and by site of death (i.e., facility vs community). Stillbirths and neonatal deaths were analyzed separately since they were thought to be due to different types of delays. All analyses were conducted using R (version 4.1.2, R Foundation for Statistical Computing).

## Results

CHAMPS enrolled 11,633 total cases during the study period; 3,231 were infants and children and were excluded, and 1,323 cases did not have sufficient information to determine if delays occurred prior to the stillbirth or neonatal death (Fig 1). Thus, 7,079 cases were included in our analyses (stillbirths, n=3,424; neonatal deaths, n=3,655). Among included stillbirths, 53.6% were male, the median gestational age was 36 weeks (IQR 32, 38 weeks), the median birth weight was 2,300 grams (IQR 1,520, 3,000 grams), and 84.9% of these stillbirths occurred in healthcare facilities (Table 1). Of the 3,424 stillbirths included in the analysis, stillbirth type was available for 2,037 cases (59.5%); among these, 1,085 (53.3%) were classified as fresh stillbirths and 952 (46.7%) as macerated.

**Fig 1.**
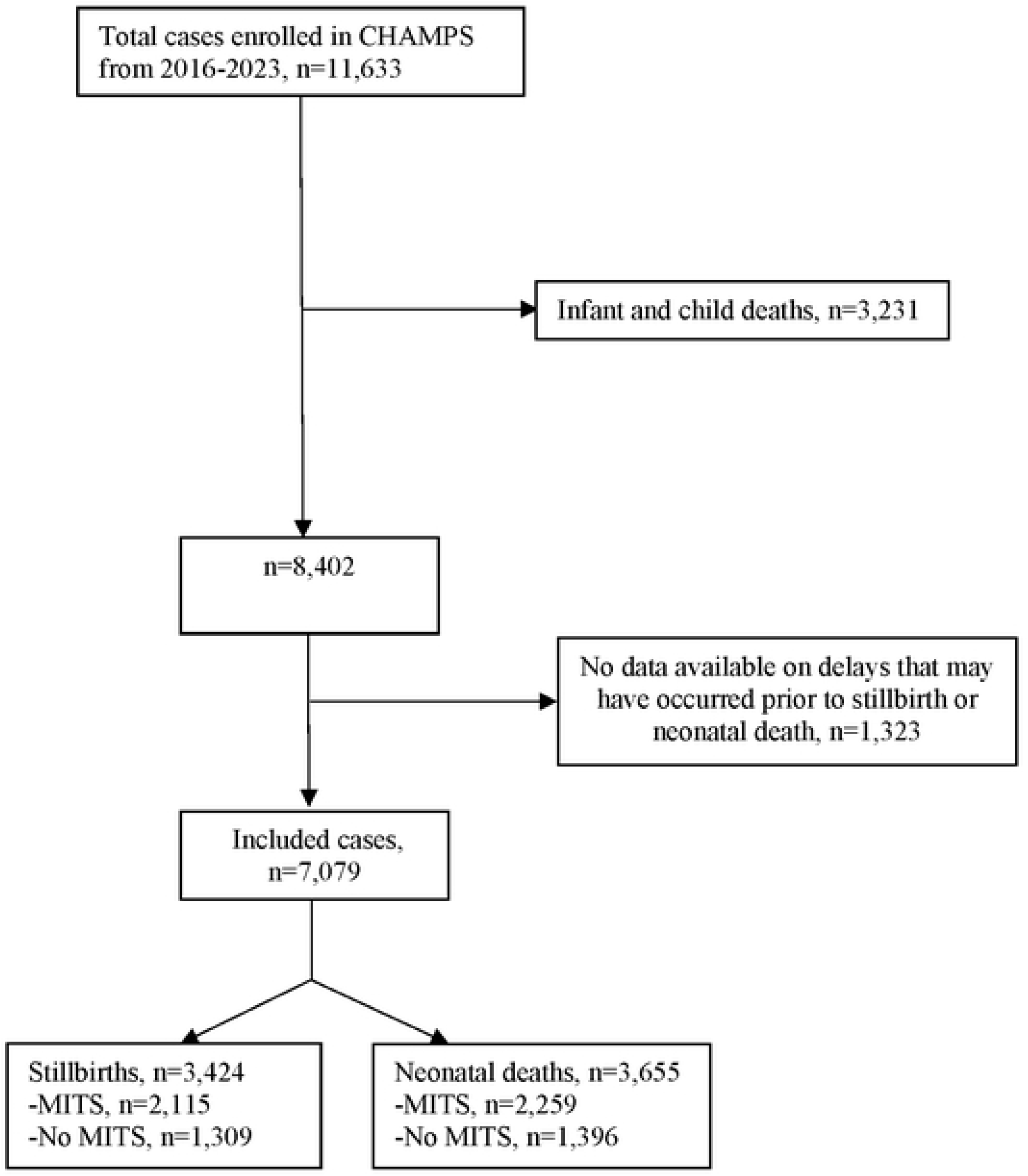
PRISMA flow diagram for stillbirths and neonatal deaths included in analyses of the “Three Delays in Healthcare” framework in the CHAMPS network, 2016-2023

**Table 1.** Description of stillbirths and neonatal deaths enrolled in the Child Health and Mortality Prevention Surveillance (CHAMPS) network.

| Characteristic | Stillbirths (N=3,424), n (%) | Neonatal deaths (N=3,655), n (%) |
| --- | --- | --- |
| **Documented maternal gestational age in weeks** |  |  |
| <28 weeks | 205 (6.0) | 446 (12.2) |
| 28-33 weeks | 469 (13.7) | 441 (12.1) |
| 34-36 weeks | 440 (12.9) | 319 (8.7) |
| 37-42 weeks | 831 (24.3) | 934 (25.6) |
| Unknown | 1,479 (43.2) | 1,515 (41.5) |
| Median, IQR | 36 [32, 38] | 34 [30, 38] |
| **Age at death in days** |  |  |
| First 24 hours | -- | 1,459 (39.9) |
| 1 to 6 days | -- | 1,494 (40.9) |
| 7 to 27 days | -- | 702 (19.2) |
| Median, IQR | -- | 2 [0, 5] |
| **Sex** |  |  |
| Male | 1,836 (53.6) | 2,095 (57.3) |
| Female | 1,564 (45.7) | 1,551 (42.4) |
| Indeterminate or Ambiguous | 5 (0.1) | 4 (0.1) |
| Unknown | 19 (0.6) | 5 (0.1) |
| **HIV status** |  |  |
| Infected | 6 (0.2) | 11 (0.3) |
| Exposed and uninfected | 191 (5.6) | 403 (11.0) |
| Uninfected or unknown | 3,227 (94.2) | 3241 (88.7) |
| **Delivery type** |  |  |
| Vaginal | 2,464 (72.0) | 2,470 (67.6) |
| C-section | 684 (20.0) | 854 (23.4) |
| Instrumentation (e.g., forceps) | 1 (0.0) | 2 (0.1) |
| Unknown | 275 (8.0) | 329 (9.0) |
| **Birth weight in grams** |  |  |
| Extremely low birth weight (<1000 grams) | 108 (3.2) | 340 (9.3) |
| Very low birth weight (1000–1499 grams) | 298 (8.7) | 454 (12.4) |
| Low birth weight (1500–2499 grams) | 629 (18.4) | 646 (17.7) |
| Normal weight (2500–4000 grams) | 765 (22.3) | 1001 (27.4) |
| Macrosomia (>4000 grams) | 51 (1.5) | 33 (0.9) |
| Unknown | 1573 (45.9) | 1181 (32.3) |
| Median, IQR (in grams) | 2,300 [1520, 3000] | 2,060 [1235, 2950] |
| **Location of birth** |  |  |
| Healthcare facility | 2,906 (84.9) | 3,029 (82.9) |
| Home | 316 (9.2) | 432 (11.8) |
| On the way to health center | 25 (0.7) | 28 (0.8) |
| Other location | 16 (0.5) | 7 (0.2) |
| Unknown | 161 (4.7) | 159 (4.4) |
| **CHAMPS site** |  |  |
| Bangladesh | 850 (24.8) | 807 (22.1) |
| Ethiopia | 639 (18.7) | 479 (13.1) |
| Kenya | 290 (8.5) | 315 (8.6) |
| Mali | 476 (13.9) | 447 (12.2) |
| Mozambique | 537 (15.7) | 684 (18.7) |
| Sierra Leone | 326 (9.5) | 343 (9.4) |
| South Africa | 306 (8.9) | 580 (15.9) |
| **MITs procedure done** | 2,115 (61.8) | 2,259 (61.8) |
| **Site of Death** |  |  |
| Healthcare facility | 3,009 (87.9) | 2,975 (81.4) |
| Community | 415 (12.1) | 680 (18.6) |
| **Hospital duration in hours, median (IQR)) (N=2778)** | -- | 28 [8, 79] |
| **Weight at MITs, in kilograms, median (IQR)) (N=4343)** | 2.1 [1.4, 2.8] | 1.9 [1.2, 2.8] |

Among neonatal deaths, 57.3% were male, the median gestational age at birth was 34 weeks (IQR 30, 38 weeks), and the median birth weight was 2,060 grams (IQR 1,235, 2,950 grams (Table 1). The majority (82.9%) of neonatal deaths were among neonates who were born in healthcare facilities, and 81.4% of the deaths occurred in healthcare facilities. The median age at the time of death among neonatal deaths was 2 days (IQR 0, 5).

### Overall Delays in the “Three Delays in Healthcare” Framework

Documented inadequate clinical care (e.g., delays in neonatal resuscitation, infrastructure limitations leading to suboptimal clinical care, etc.) was the most identified delay for stillbirths (Fig 2). Family under-estimation of the neonate’s illness severity was the most common delay among neonatal deaths. Overall, 96.8% (n=3,313) of stillbirths and 97.7% (n=3,571) of neonatal deaths had ≥1 delay identified (S1 Table and S2 Table). There were 240 (7.0%) stillbirths and 1,389 (38.0%) neonatal deaths that had ≥1 delay identified in all three categories in the “Three Delays in Healthcare” framework.

**Fig 2.**
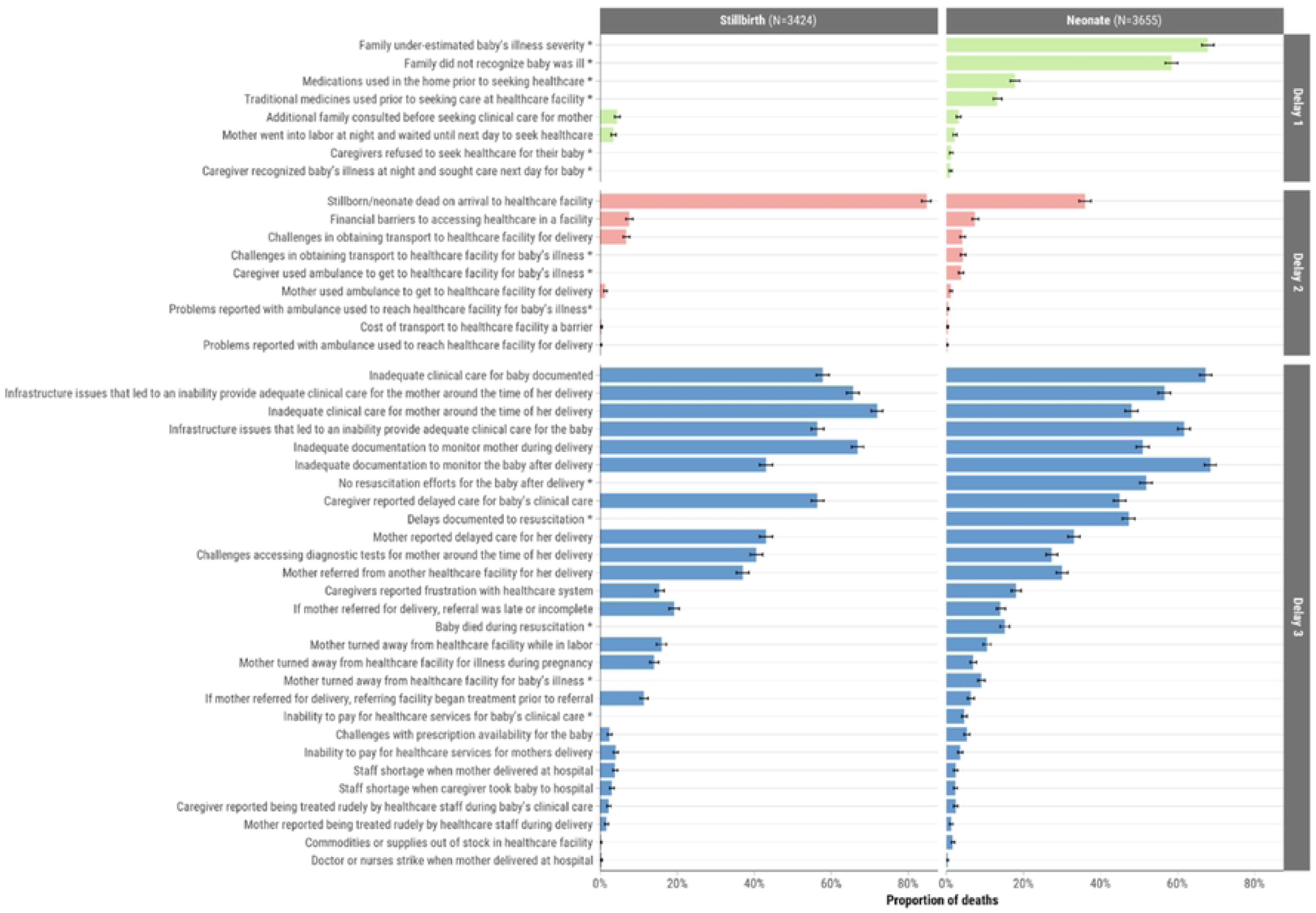
Proportion of included stillbirths and neonatal deaths that experienced each type of delay in the “Three Delays-in-Healthcare” framework

The most common delays in neonatal deaths varied by the time of death (S1 Figure). For example, delays in the home (e.g., under-estimation of illness risk) was more common among neonates who died at age 7 to 27 days and delays in quality clinical care (e.g., inadequate monitoring of mother during delivery and of baby after delivery) were more common among neonates who died in the first 24 hours of life. The proportion of cases that had at least one delay did not vary substantially by year (S2 Figure).

Delays were not more common among male stillbirths compared to female stillbirths (S1 Table), or among male neonatal deaths compared to female neonatal deaths (S2 Table). Delays in illness recognition and in reaching a healthcare facility were more common among stillbirths and neonatal deaths that occurred in the community than among those that occurred in healthcare facilities (S1 Table and S2 Table).

### Site Variation in Delays in the “Three Delays in Healthcare” Framework

The proportion of stillbirths and neonates who experienced delays before death varied by site (Fig 3). Overall, delays in illness recognition, reaching a healthcare facility, and the provision of clinical care were most common among stillbirths and neonatal deaths that were enrolled in Bangladesh (*P*<0.001 compared to other sites; S1 Table and S2 Table). Delays in the provision of healthcare were least common for stillbirths and neonatal deaths in South Africa (S3 Fig). There was no substantial difference in delays by sex of the deceased and site (Fig 3).

**Fig 3.**
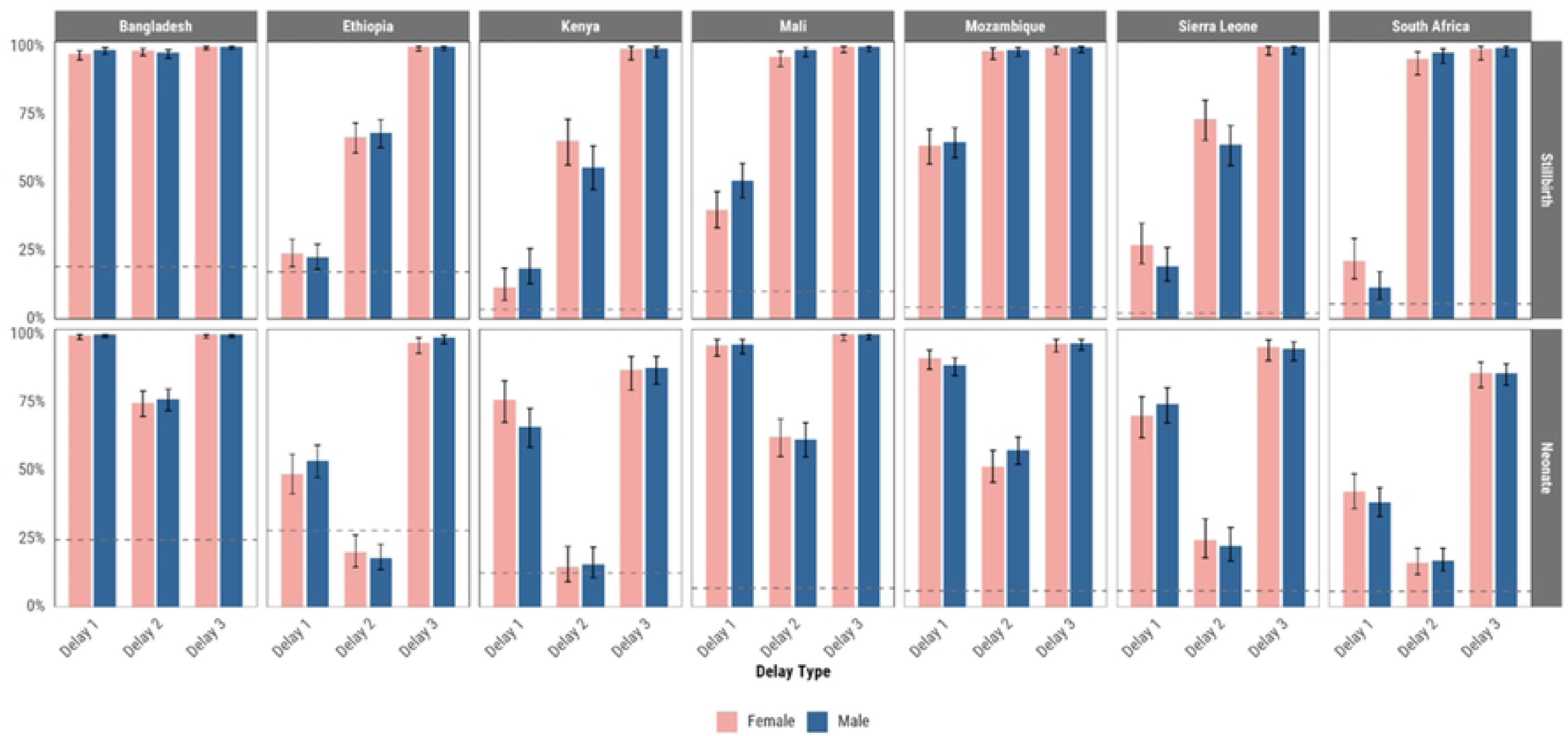
Proportion of stillbirths and neonates who died and experienced each delay in the “Three Delays-in-Healthcare” framework by site and sex.* **\***Bars represent the proportion of deaths with at least one delay identified within each category. Error bars indicate 95% confidence intervals. Dashed horizontal lines indicate the proportion of deaths that occurred following home deliveries in each site and age group.

### Variation in Delays by Cause of Death for Stillbirths and Neonatal Deaths

Among the 2,115 stillbirths who underwent the full postmortem determination of cause of death procedures, the most common causes of stillbirth anywhere in the causal chain were intrauterine asphyxia/hypoxia (n=1696, 80.2%), congenital birth defects (n=184, 8.7%), and sepsis (n=109, 5.2%). Among stillbirths, the most identified maternal conditions were maternal hypertension (n=2,718, 79.4%), placental complications (n=2,094, 61.2%), and anemia (n=1,570, 45.9%).

Among the 2,259 neonates who underwent the full postmortem determination of cause of death procedures, the most common causes of death anywhere in the causal chain were preterm birth complications (n=933, 44.1%), perinatal asphyxia/hypoxia (n=859, 40.6%), and sepsis (n=798, 37.7%). The most common maternal conditions in neonatal deaths were hypertension (n=1,901, 52.0%), multiple gestation (n=1,370, 37.5%), and placental complications (n=1,168, 32.0%). Types of delays did not differ substantially by maternal condition (Fig 4). There was no substantial variation in the specific delays experienced by stillbirths (S4 Fig) or neonates (S5 Fig) when grouped by causes of death.

**Fig 4.**
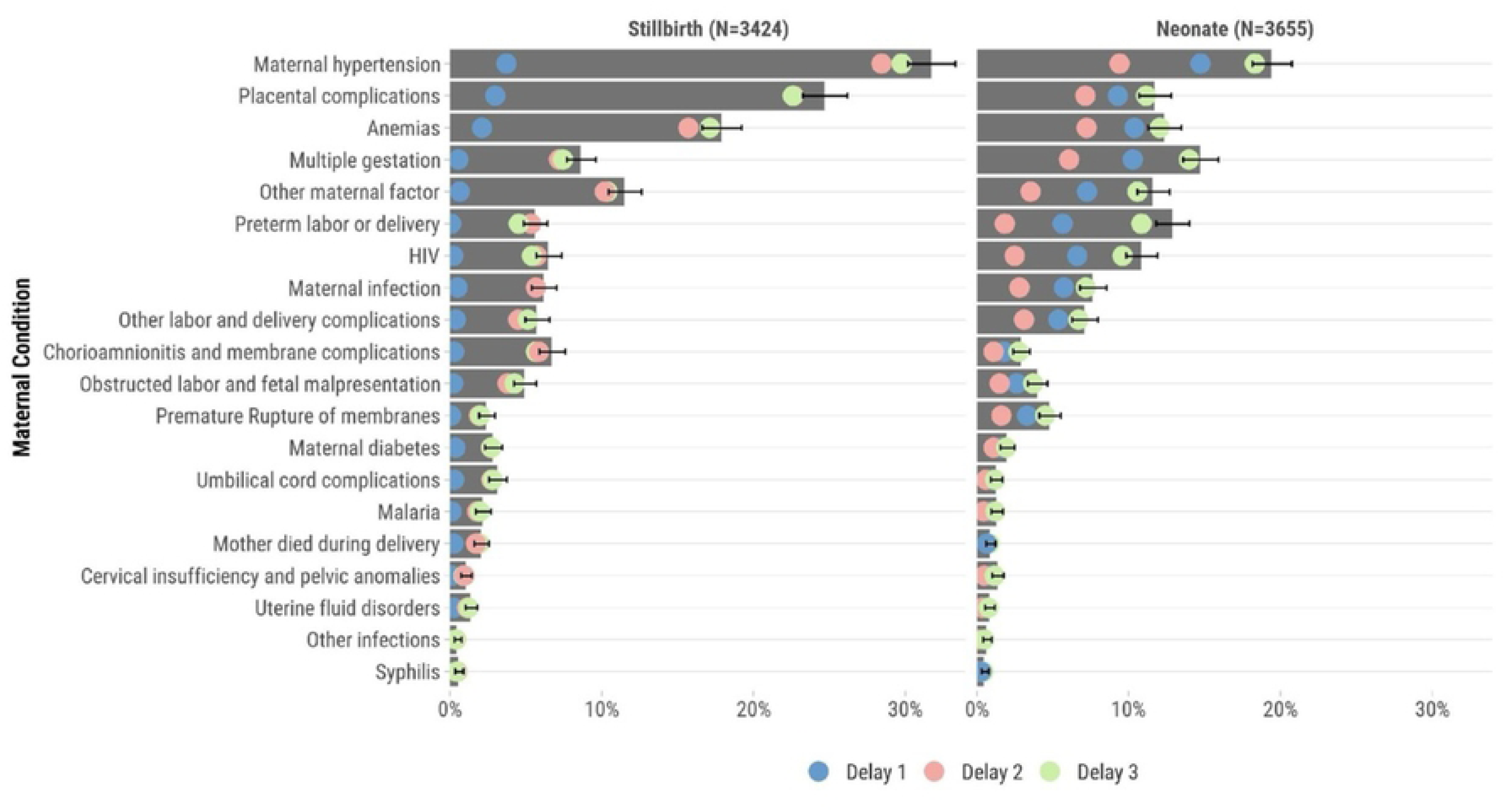
Maternal conditions and delays among a) stillbirths and b) neonatal deaths* **\***Bars are proportions of stillbirths/neonates with each maternal condition and dots are proportions of each delay relative to the bar length

### Antenatal Care for Stillbirths and Neonatal Deaths

Among the 3,424 stillbirths, 73.7% of mothers had ≥1 antenatal care visit documented and only 66.3% of the 3,655 mothers whose neonate died had ≥1 antenatal care visit documented (Fig 5). Only 54 (1.6%) stillbirths and 33 (0.9%) neonatal deaths had documentation of eight or more antenatal care visits, as recommended by the WHO.^25^ The median number of antenatal care visits was 2 (IQR 1, 4) for both stillbirths and neonatal deaths. Antenatal care visits were least common during the first trimester and most common during the third trimester of pregnancy for both stillbirths and neonatal deaths. The proportion of included cases with any documented antenatal care visits was lowest among cases enrolled at the Ethiopia site and highest among cases enrolled at the Bangladesh site (S6 Fig).

**Fig 5.**
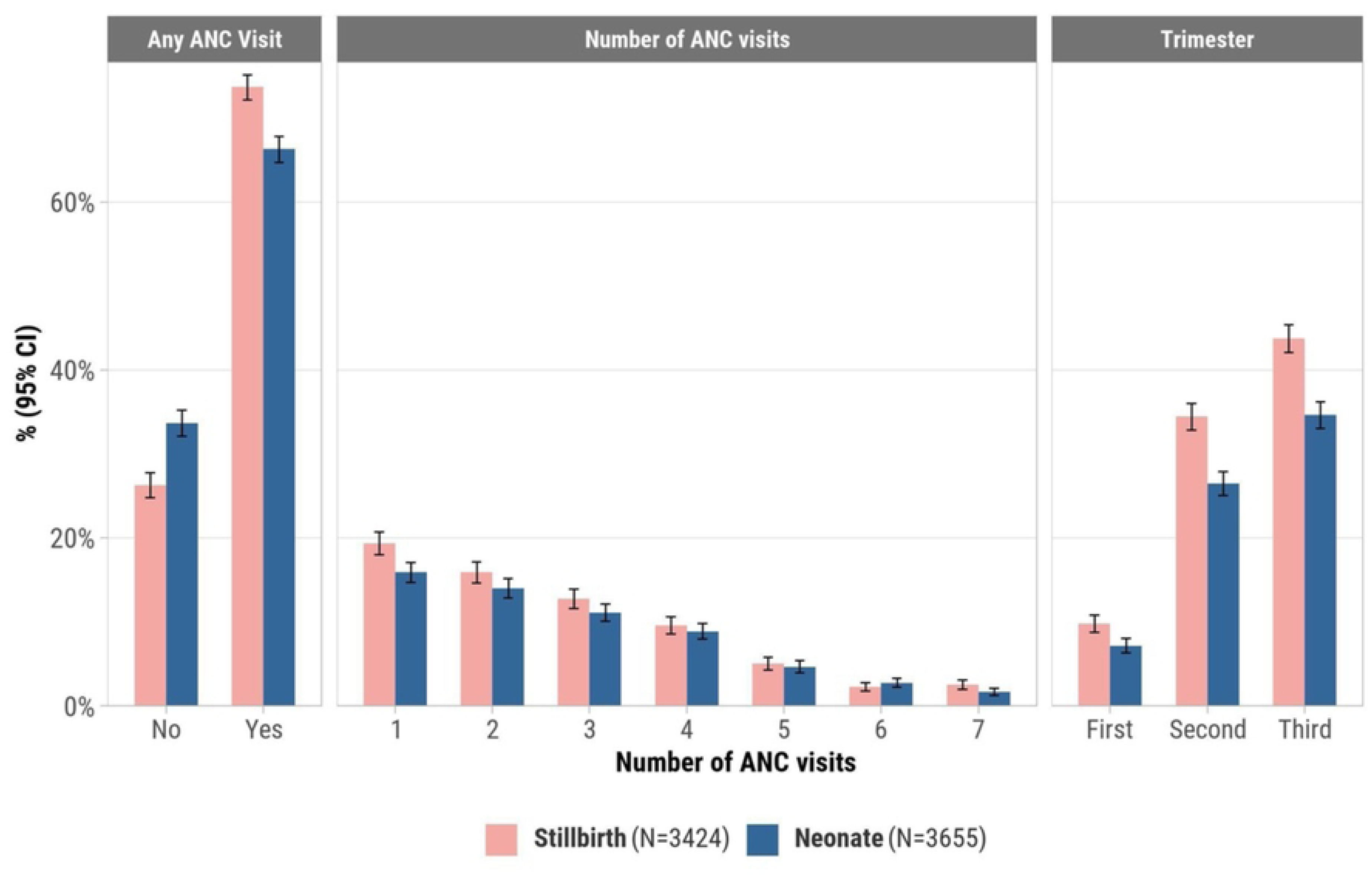
Distribution and timing of antenatal care visits for mothers of stillbirths (N=3,424) and neonates who died (N=3,655)

## Discussion

In our study of 3,424 stillbirths and 3,655 neonatal deaths in regions with high rates of childhood mortality in seven low- and middle-income countries, delays in the decision to seek care, delays in reaching healthcare facilities, and delays in the provision of healthcare before death occurred in almost all cases. Delays occurring in healthcare facilities were most common. Although all three delays were common across the CHAMPS network, specific types of delays varied by site. The WHO encourages eight antenatal care visits during pregnancy as there is evidence associating frequent antenatal care visits with reduced perinatal deaths and improved maternal outcomes; we observed that antenatal care among mothers of stillbirths and neonatal deaths was infrequent and often began late in pregnancy.^25^

Prior studies in the CHAMPS network have demonstrated that the local expert review panels who review each death frequently note that delayed access to, and suboptimal, clinical care are the most common ways in which stillbirths and neonatal deaths could have been averted.^26^ Also consistent with previous studies conducted in Kenya,^27^ we found that delayed provision of high quality clinical care was the most common type of delay experienced by stillbirths and neonatal deaths. Prior modeling studies suggest that health service strengthening may have the largest impact on neonatal mortality reduction in low- and middle-income countries.^28^ Furthermore, results from a previous systematic review suggest that common delays related to maternal and newborn care in healthcare facilities include lack of appropriate storage for medications, lack of equipment, and limited surgical, transfusion and laboratory facilities are common delays in healthcare facilities in sub-Saharan Africa and South Asia.^29^ Thus, additional efforts to enhance health services available to pregnant mothers both during pregnancy to enhance the detection of maternal conditions and around the time of delivery and to neonates are warranted.^30^

Our findings also suggest that a site-specific tailored approach to health services strengthening may be indicated. Resources needed to monitor mothers during delivery and equipment for neonatal resuscitation vary in sub-Saharan Africa and Bangladesh and even within countries. Thus, an assessment of needed resources should be conducted at the site level.^28^ The site variation that we observed suggests that interventions tailored at illness recognition for mothers may be better suited for some CHAMPS sites, while enhanced healthcare provision may be most needed in other CHAMPS sites. Ultimately, additional studies are warranted to better elucidate reasons for delayed healthcare provision among stillbirths and neonatal deaths.

In our study, <10% of mothers of stillbirths and neonates who died had their first antenatal care visit in the first 12 weeks of pregnancy and none had the recommended eight antenatal care visits. Because frequent antenatal care visits may enhance timely detection and management of potential pregnancy-related complications, improve communication between healthcare providers and pregnant women, and may increase the likelihood of positive pregnancy outcomes, the WHO recommends that the first antenatal care visit occur before the 12^th^ week of pregnancy and that expecting mothers have at least eight antenatal care visits during pregnancy.^25^ Meta-analysis level data suggest that antenatal care visits in sub-Saharan Africa reduce the risk of neonatal mortality by as much as 39%.^31^ Moreover, results from one study in Ethiopia suggest that high-quality antenatal care may reduce the odds of a mother having a stillbirth by as much as 80%.^32^ However, other studies in Bangladesh suggest no differences in perinatal outcomes for mothers who had more antenatal care visits,^33^ though the quality of care provided in such visits was less clear. Thus, additional efforts may be needed to augment both the frequency of and quality of antenatal care visits in settings like those in which CHAMPS operates.

### Limitations

The results of our multi-country study should be interpreted in the context of certain limitations. Given the observational nature of our study, we cannot infer any causality between the delays observed and either the stillbirth or the death of the neonate. Furthermore, it is possible that there were additional, unmeasured delays among >7,000 stillbirths and neonatal deaths. Thus, our estimates of delays in healthcare seeking and provision may be an underestimate of the frequency of such occurrences. We could not compare the frequency of antenatal care visits among mothers of stillbirths and neonates who died to the frequency of such visits among neonates who survived. Illness recognition and decisions to seek healthcare are often influenced by culture and societal norms,^34,35^ which we could not measure in our study. In many cases, the specific elements of clinical care that were delayed were not recorded. Thus, additional studies are warranted to understand specific delays in clinical care administration. We could not determine if maternal diagnoses were missed during antenatal care visits and the potential contribution of such missed diagnoses to stillbirths or neonatal deaths. We used a rigorously validated large language model to extract discrete variables from narrative fields, which may have introduced some assumptions made by the model. We overcame this issue by iteratively training the model on the specific study questions and by manually reviewing a large proportion of cases to ensure that the model did not hallucinate or make inferences that should not have been made regarding delays experienced before the stillbirth or death of the neonate.

## Conclusions

Delayed illness recognition and healthcare seeking by caregivers and delayed healthcare provision were common among stillbirths and neonatal deaths in seven low- and middle-income countries. Additional efforts are warranted to promote early and complete antenatal care among mothers. Despite high facility delivery rates, healthcare provision delays were the most predominant, suggesting that strengthening facility-based clinical care quality, rather than simply increasing facility access, may be the most impactful intervention for reducing perinatal mortality in these settings.

## Data Availability

CHAMPS data are available through the CHAMPS website: https://champshealth.org/causes-of-death/ Additionally, specific data requests can be made through: https://champs.emory.edu/redcap/surveys/?s=PCEERX993Y

https://champshealth.org/causes-of-death/

## Acknowledgements

We extend our deepest gratitude to the families who enrolled in CHAMPS to understand why their child died. We would also like to thank the CHAMPS staff in each site for their insights, expertise, and continued commitment to ensuring that high quality data are collected in this project and available globally for developing strategies and interventions to save lives.

## Abbreviations

CHAMPS: Child Health and Mortality Prevention Surveillance
MITS: minimally invasive tissue sampling
DeCoDe: Determination of Cause of Death
LMIC: low- and middle-income country
IQR: interquartile range
WHO: World Health Organization

## Supplementary Information

**S1 Table**. Proportion of stillbirths that experienced each delay in the “Three Delays-in-Healthcare” framework by site, sex, and site of death (N=3,424)

**S2 Table**. Proportion of neonatal deaths that experienced each delay in the “Three Delays-in-Healthcare” framework by site, sex, and site of death (N=3,655)

**S1 Fig**. Proportion of included neonatal deaths that experienced each type of delay in the “Three Delays-in-Healthcare” framework by age of neonate at the time of death

**S2 Fig**. Proportions of included cases that experienced each delay over the study period

**S3 Fig**. Proportion of delays in the “Three Delays-in-Healthcare” framework among deceased a) stillbirths (N=3424 and b) neonatal deaths (N=3655) by country site

**S4 Fig**. Frequencies in delays in the “Three Delays in Healthcare” framework for the leading causes of stillbirths (N=2,115)

**S5 Fig**. Frequencies in delays in the “Three Delays in Healthcare” framework for the leading causes of neonatal deaths (N=2,259)

**S6 Fig.** Proportion of enrolled cases with antenatal care (ANC) visits documented by CHAMPS site

